# A QUANTITATIVE ASSESSMENT OF VISUAL FUNCTION FOR YOUNG AND MEDICALLY COMPLEX CHILDREN WITH CEREBRAL VISUAL IMPAIRMENT: DEVELOPMENT AND INTER-RATER RELIABILITY

**DOI:** 10.1101/2024.05.07.24306570

**Authors:** Kathleen M. Weden, Elizabeth A. Barstow, Robert A. Oster, Dawn K. DeCarlo

## Abstract

**Background:** Cerebral Visual Impairment (CVI) is the most common cause of low vision in children. Standardized, quantifiable measures of visual function are needed.

**Objective:** This study developed and evaluated a new method for quantifying visual function in young and medically complex children with CVI using remote videoconferencing.

**Methods:** Children diagnosed with CVI who had been unable to complete clinic-based recognition acuity tests were recruited from a low-vision rehabilitation clinic(n=22)Video-based Visual Function Assessment (VFA) was implemented using videoconference technology. Three low-vision rehabilitation clinicians independently scored recordings of each child’s VFA. Interclass correlations for inter-rater reliability was analyzed using intraclass correlations (ICC). Correlations were estimated between the video-based VFA scores and both clinically obtained acuity measures and children’s cognitive age equivalence.

**Results:** Inter-rater reliability was analyzed using intraclass correlations (ICC). Correlations were estimated between the VFA scores, clinically obtained acuity measures, and cognitive age equivalence. ICCs showed good agreement (ICC and 95% CI 0.835 (0.701-0.916)) on VFA scores across raters and agreement was comparable to that from previous, similar studies. VFA scores strongly correlated (r= -0.706, p=0.002) with clinically obtained acuity measures. VFA scores and the cognitive age equivalence were moderately correlated (r= 0.518, p=0.005), with notable variation in VFA scores for participants below a ten month cognitive age-equivalence. The variability in VFA scores among children with lowest cognitive age-equivalence may have been an artifact of the study’s scoring method, or may represent existent variability in visual function for children with the lowest cognitive age-equivalence.

**Conclusions:** Our new VFA is a reliable, quantitative measure of visual function for young and medically complex children with CVI. Future study of the VFA intrarater reliability and validity is warranted.

## INTRODUCTION

Children with developmental delay are at a higher risk of visual impairment^1^ and one of the most common causes of child vision impairment is Cerebral Visual Impairment (CVI).^2^ CVI is defined as a verifiable visual dysfunction which cannot be attributed to disorders of the anterior pathways or any potentially co-occurring ocular impairment ^3^. Pediatric CVI is a diagnosis of exclusion derived from medical history, neuroimaging, visual function examination, and caregiver interview.^4^ While some spontaneous improvement in visual function over time has been documented in children with CVI,^5^ these children usually remain visually impaired and have decreased global functioning and poorer developmental outcomes.^6^ Functional outcomes have been shown to significantly improve for children with CVI when intervention is provided at a young age,^7^ making delay in diagnosis detrimental to quality of life across the life span.^8^ Therefore, there is a need for earlier and increased identification and better assessment of CVI.^9^

### Assessment of Pediatric CVI

There is no standard protocol for assessing visual function in pediatric CVI and vision assessment is complicated by multiple deficits across domains of function (i.e.: impairment in one or multiple visual functions, including visual acuity, contrast sensitivity, color recognition, oculomotor functions (pursuits, saccades), and visual fields;^10^ and impairment in higher-order visual perceptual functions such object identification, visuospatial processing, visual attention and visually guided reach).^4,11^ Therefore, clinical assessment typically includes examination of ocular function, preferential-looking tests of acuity and contrast sensitivity, and structured history-taking and clinical observation.^12^ Additionally, CVI can present varying degrees of deficit across different visual functions,^13^ and there are few standardized methods for quantifying these variations in visual functions for this population.

Most children with CVI have some intact visual function, but may have higher-order visual processing challenges such as difficulty with visually complex environments, facial recognition, and poor visual regard of novel items.^14^ There are also visual behaviors that are particularly common among young children and babies with CVI, including preferred attention to objects with movement, visual latency, difficulty with distance viewing, and difficulty with visual complexity.^15^

To address the need for standardized, quantifiable measures of visual function and visual perception for children diagnosed or at risk for CVI, computer-based activities with integrated eye-tracking systems have been trialed.^16–18^ However, these computer-based activities have been developed for children with medium to high levels of visual ability and higher cognitive levels. Screen-based methods for measuring acuity in young children using a preferential-looking paradigm have also been trialed and have shown high correlation with gold-standard clinic measures.^19,20^ But these studies did not target medically complex children at lower levels of cognition or visual function.

In one study a video-based assessment was developed to quantify oculomotor and visual function in young children with low vision, who also had cognitive and communication impairment.^21^ The study demonstrated that a video-based functional vision assessment strongly correlated with subjective clinical assessment of fixation and smooth pursuit. It also demonstrated success of a video-based method for quantitative assessment of visual functions for young and medically complex children with general vision impairment. Further investigation is warranted to understand the success of a video-based modality for young and medically complex children with CVI.

### Study Purpose

The purpose of this study was to develop a Video-based Visual Function Assessment (VFA) for young and medically complex children with CVI and examine the inter-rater reliability among clinician raters. Research questions were: what is the inter-rater reliability of the VFA; are there differences in reliability between the different videos used for the VFA; are there attributes of the videos that produce higher rates of reliability; and how do clinical characteristics such as acuity and cognitive age relate to VFA scores?

## METHODS

This study used a within-subjects design, where each participant was assessed for all outcome measures and in the same order. There were three outcome measures: mean total VFA score per rater, visual acuity, and cognitive age equivalent. All measures were obtained sequentially on the same day, in a clinical exam room within the [name withheld], an outpatient clinic located within [name withheld]. Assessments were done between October, 2022 and March, 2023.

### Participants

The study inclusion criteria were: diagnosis of CVI, 0-18 years chronological age, English-speaking, and no history of completing a recognition acuity test such as a Snellen test.

Recruitment occurred either during a regular clinical exam, or with a letter and follow-up phone call invitation. Consent was completed prior to assessment with the VFA. Caregivers received ten dollars and free parking upon completing the in-clinic assessments, and an additional ten dollars after they completed the follow-up interview.

### VFA Development

The VFA was developed through an iterative process of trial and redesign. Foundational visual function skills of fixation and smooth pursuit were assessed, as well as other factors known to support visual function for children with CVI (e.g., limiting clutter, using highly saturated colors, limiting environmental distraction or moving targets, and sustained presentation of stimuli^15,22^).

In its final form, the VFA was comprised of three different videos of two to three minutes:

- Video One: large static dots (angular subtense of 18.08 degrees) with no competing stimuli; dot color alternated red and yellow and randomized to appear in one quadrant. Half of video used solid black background and half used solid white background. Two-dimensional “Elmo” displayed randomly between stimuli with accompanying song clip.
- Video Two: spinning cross (angular subtense of 18.59 degrees) with no competing stimuli; cross alternated between solid black or white; and randomized to appear in one quadrant. Background was a solid contrast in black or white. Two-dimensional video clips of saturated dancing fruit appeared randomly between stimuli with an accompanying song clip.
- Video Three: modeled after the Teller Acuity Cards II, a square box with gratings (angular subtense of 19.59 degrees) was presented randomly to the right or left side of the screen. Each of the three gratings (0.43 cycles-per-centimeter (cpcm), 1.6 cpcm, 6.5 cpcm) was presented three times. To regain attention to the center of the screen, a static shape (alternating between star, heart, or lightening bolt) of saturated solid color (red, yellow, blue) appeared centrally on the screen after each grating. Three short 2D video clips of saturated dancing objects were interspersed with accompanying sound clip.

While Video One and Video Two both presented stimuli for testing fixation and smooth pursuit, the types of stimuli differed between the videos (See Figure 1). Our aim with Video Three was to conduct a preliminary trial for testing acuity with this population using a video-based format.

**Figure 1.**
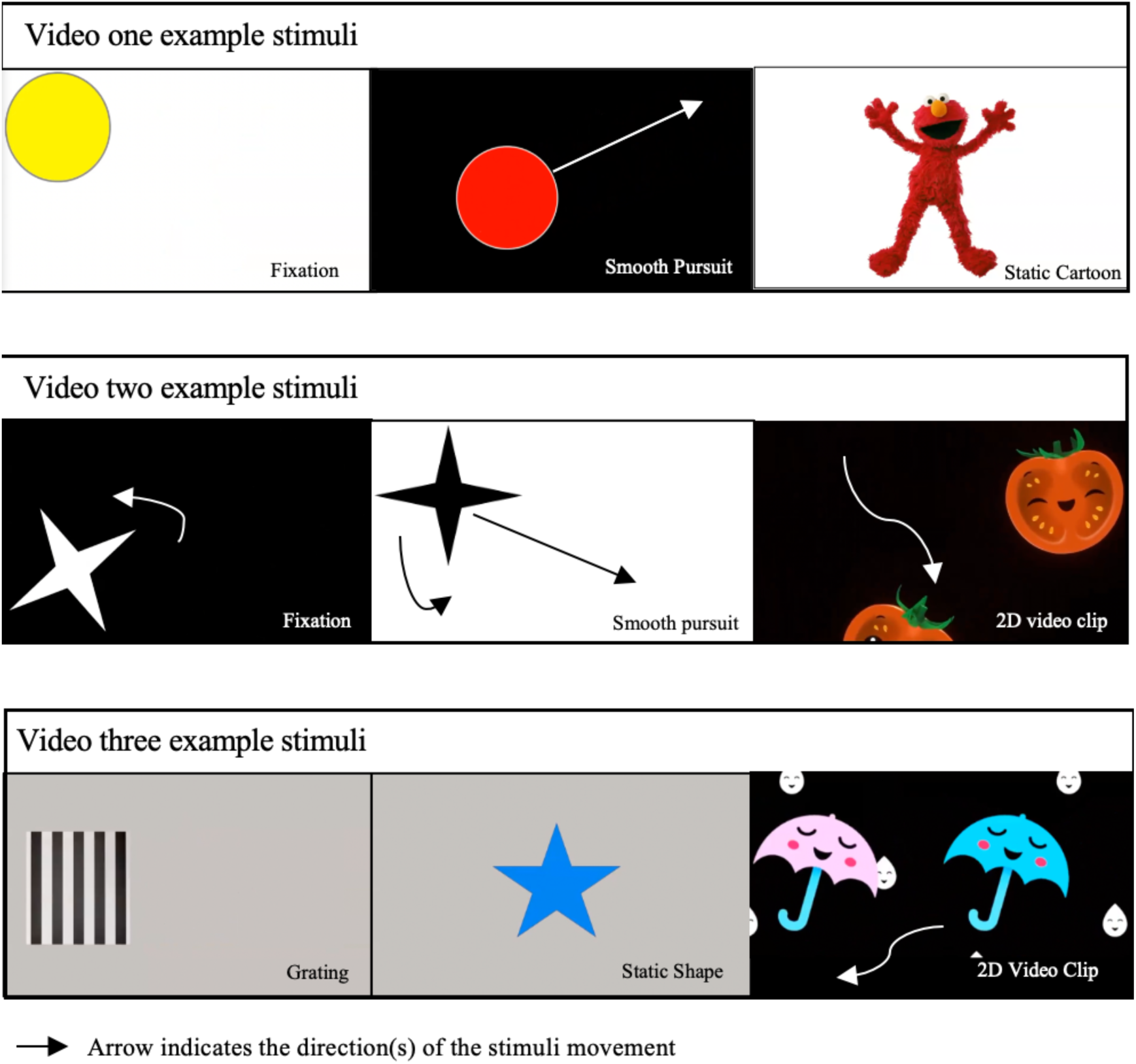
Still image examples of the stimuli used in the Video-based Visual Function Assessment (VFA) Videos.

Then if the VFA appeared successful in assessing fixation and smooth pursuits with this population, full assessment of visual acuity would be the next visual function we would aim to add to the protocol. For this initial trial we decided to abbreviate the number of gratings presented to minimize the total assessment time for the participants.

Stimuli presentation time was four seconds for static stimuli and eight seconds for smooth pursuit stimuli, similar to that of other video-based vision assessments for children with CVI.^19,23^ Various short sound bites were included between static stimuli, and short high-contrast two-dimensional video clips were used between tracking stimuli to encourage the child’s attention. The VFA was strategically built as three separate videos so that breaks could be offered between videos.

### Setup

The VFA assessment was setup in a research room within the [name withheld], an outpatient clinic located within [name withheld]. A 34-inch computer monitor, with 3440 by 1440-pixel resolution, was mounted onto an adjustable arm and attached to a mobile cart. A standard video-conference camera was used to capture video of the child during the Zoom-based video-conference assessment. The camera was mounted on the bottom edge, centered on the monitor.

Both overhead and room lighting were turned off during the assessment. In two cases the darkened room was not tolerable for the child and so overhead lighting remained. A dimmable LED light was placed on both sides of the child, at shoulder height to remove shadowing around the orbit and bridge of the child’s nose.

The child was placed in a seated position of their preference (e.g., in their wheelchair, a stroller or adaptive stroller, on a caregiver’s lap, or independently on a supportive chair), directly in front of the monitor at 55cm, measured from the camera to the child’s nose. The child was positioned upright as close to 90 degrees as possible. When head control was limited, posterior or lateral supports were added around the headrest, or the caregiver provided hands-on support. The VFA was projected through a live videoconference using the Zoom platform. After setup with the researcher, the child and caregiver remained in the room while the researcher moved to an adjacent room, and the VFA was conducted as a mock-remote assessment. Caregivers were cued before and occasionally during the assessment through the video conference to adjust their child’s head positioning. In collaboration with the researcher administering the assessment, the caregivers supported or facilitated their child in a way they knew to be most effective.

Recordings of all three videos were captured through the Zoom program. Within the program, the child’s image was mirrored so that their gaze matched the direction of the stimuli as it appeared to the assessment administrator. The child’s image was also maximized within the Zoom program to facilitate observation of their pupils when rating from the video recording.

### Measurement Protocol

The study protocol included three measures: the VFA, cognitive assessment with the Bayley III Cognitive Subtest,^24^ and acuity testing with the Teller Acuity Cards II.^25^ The protocol was designed to minimize the burden for the child and their family, and assessments were discontinued at the caregiver’s discretion if the child was determined to be agitated. Thus, assessment with the VFA was always performed first, followed by developmental testing with the Bailey III, and lastly the Teller Acuity Card test. The administration of these measures was conducted and modified according to the child’s tolerance, as detailed below.

#### Bayley III Cognitive Subtest

The Bayley III Cognitive Subtest was selected for a few reasons. First, it was preferable to assess the child the day they were in the clinic, rather than use a more general inventory such as the Pediatric Evaluation of Disability Inventory (PEDI),^26^ because the abilities of children with CVI can fluctuate from day to day. Second, the cognitive subtest can be administered on its own, and this minimized testing time for our participants. The Bayley III has been revised to be less reliant on the child’s receptive language skills and motor ability, and also details accommodations and adaptations that can be used for children with sensory and physical impairments. It can also be used with children of an older chronological age who are developmentally delayed, which fit our convenience sample. Norm-referenced scores are not valid when used with this type of population, instead the developmental age equivalent is to be used and cognitive age equivalent was meaningful for the purposes of this study.

#### Teller Acuity Cards II

Many of our recruited participants came directly from clinic visit, during which the optometrist had often conducted either Teller Acuity Cards II, Lea Acuity Paddle testing, or the HOTV forced choice optotype acuity testing and the prior obtained clinical measure was used. For participants unable to complete acuity testing the day of VFA assessment, their chart was reviewed and any recorded acuity from the past year was used.

### Scoring of VFA

Prior to rating participants from video recordings, a scale from zero to five was developed. Description was provided for each of the six conditions of looking behavior, (See Appendix A). For each stimulus presented, a score was assigned to indicate the degree of fixation or smooth pursuit that the rater observed from the child’s looking behavior. A zero-score indicated no purposeful gaze upon the stimuli and a score of five indicated a discrete and consistent gaze upon the stimuli. All three video recordings for all participants were scored independently by three raters. One rater is an occupational therapist (OT) with pediatric inpatient rehabilitation and early intervention experience, the second rater is an OT and PhD with AOTA certification in low vision rehabilitation, and the third rater is an optometrist and PhD with residency training in vision rehabilitation and an expert in pediatric low vision rehabilitation.

### Data Analysis

All acuity measures (grating and optotype) were converted from cy/cm or Snellen to logMAR. It was determined that the six-level scoring system, which offered ordinal data, limited our precision in deriving total VFA scores. With the ordinal data we would need to use either a mode or median of the individual scores per stimuli, and in doing this we would lose some of the specificity that we were hoping to gain by offering multiple standardized stimuli presentations. Therefore, all individual scores were converted from a six-level score to a zero or one score.

Using the descriptions outlined in our manual, it was determined clinically appropriate to convert any zero, one or two score to a zero, indicating impaired or absent visual function (fixation or smooth pursuit). We converted all three, four or five scores to a one, indicating intact or present visual function.

Total scores were then determined per participant and per rater for Video One and Video Two by taking the average of all rated stimuli by rater. A final acuity estimate was derived from Video Three by determining the highest grating (from 0.43 cy/cm, 1.6 cy/cm, and 6.5cy/cm) for which the child showed discrete fixation (a score of 3,4, or 5) in at least two of the three presentations. Total scores for fixation were derived by averaging the scores for all static stimuli presented in Video One and Video Two, and a total score for smooth pursuits was derived by averaging the scores for all smooth pursuit stimuli presented in Video One and Video Two. A similar method was used to determine total scores for other assessment attributes (black background and white background).

To understand the significance of differences between participants’ chronological age and their Bayley III cognitive subtest age equivalencies, a paired t-test was used. Intraclass correlations (ICCs) and their 95% confidence intervals were obtained as measures of agreement of the three raters for the video scores and for the individual video characteristics (fixation, smooth pursuits, black background, white background) of the videos. ICCs were assessed according to previously established guidelines.^27^

The means of the three raters’ scores for video recordings and sub-groupings were used in subsequent (Pearson) correlation analysis and linear regression analysis, where the VFA mean score served as the dependent variable in the linear regression analysis. Regression and correlation coefficients (r) were tested for statistical significance, and the coefficient of determination (R^2^) was used as a goodness-of-fit measure for the regression models. For the mean scores, normality of distribution was confirmed using box plots, stem and leaf plots, and normal probability plots, and the Kolmogorov-Smirnov test. Statistical tests were two-tailed, and statistical significance was set at p < 0.05. SAS software was used to perform the statistical analyses, including obtaining the ICCs. (version 9.4; SAS Institute, Cary, NC) Data were stored in a REDCap server (Research Electronic Data Capture)^28^ hosted by [name withheld]. Ethical approval for this research was obtained from [name withheld] Institutional Review Board.

## RESULTS

We prospectively recruited 24 children for the in-clinic protocol, but two VFA recordings were lost to technical failures with the Zoom platform or with camera connectivity. Descriptive statistics characterize the demographic and clinical profile of the participants (n = 22; See Table 1). Among our participants, the most common medical conditions were seizure (59.1%), prematurity (59.1%), and brain injury (22.7%), and 81.8% had multiple medical conditions that could be determinants of CVI. Cerebral palsy was a frequent co-morbidity (59.1%), and 100% were reported by caregivers to have developmental delays. There was a statistically significant difference between chronological age and cognitive age equivalent (t= 8.8, p< 0.001).

**Table 1.**
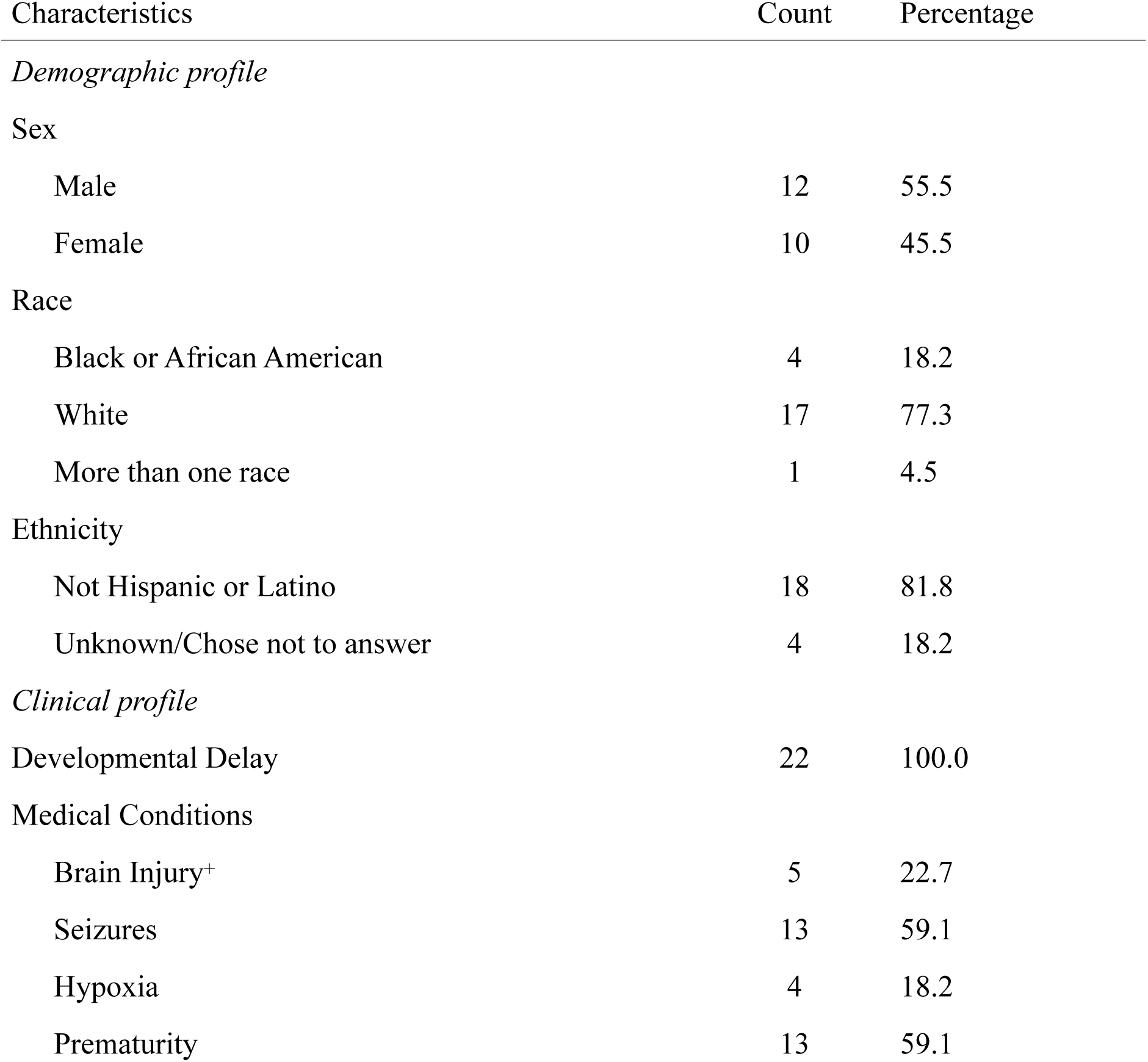
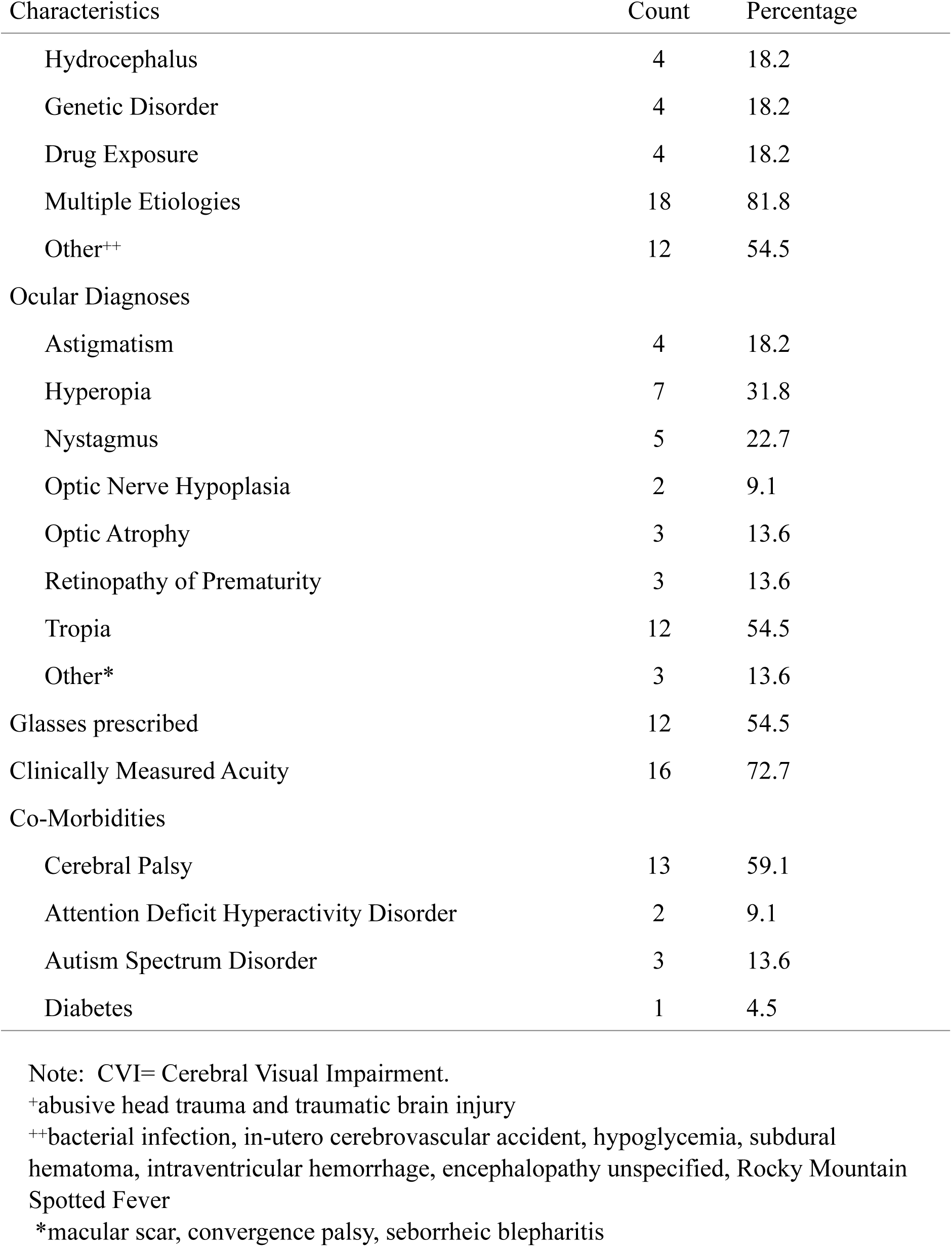
Descriptive statistics for children with CVI participating in the study.

The mean score of Video One and Video Two combined was 0.411 with a standard deviation of 0.288 and a range of 0.012 to 0.838 (See Table 2). A small percentage of participants received total average scores less than 0.10 (i.e., from from Video One n = 3 or 13.6%, and from Video Two n = 4 or 18.1%).

**Table 2.** Summary statistics for total VFA scores by video or video stimuli sub-group.

| Video or video stimuli sub-group | Summary statistics |  |  | Intra-class correlation |  | Number of total VFA scores |
| --- | --- | --- | --- | --- | --- | --- |
|  | mean | standard deviation | range | ICC | (95% CI) |  |
| Video one | 0.392 | 0.267 | 0.020-0.833 | 0.835* | (0.701, 0.916) | 66 |
| Video two | 0.442 | 0.338 | 0.000-0.967 | 0.762* | (0.591, 0.877) | 66 |
| Video one and two combined | 0.411 | 0.288 | 0.012-0.838 | 0.861* | (0.743, 0.930) | 66 |
| Video stimuli sub-groups |  |  |  |  |  |  |
| Fixation | 0.475 | 0.311 | 0.016-0.952 | 0.848* | (0.723, 0.923) | 66 |
| Smooth pursuits | 0.283 | 0.276 | 0.000-0.872 | 0.679+ | (0.478, 0.830) | 66 |
| Black background | 0.392 | 0.266 | 0.019-0.833 | 0.762* | (0.591, 0.877) | 66 |
| White background | 0.388 | 0.296 | 0.000-0.863 | 0.819* | (0.676, 0.908) | 66 |

Abbreviations: VFA= Visual Function Assessment. ICC= Intra-class correlation coefficient. CI= Confidence interval.
Notes: Summary statistics for the total VFA scores are calculated from scores made by three clinical raters who scored every child (n=22) on video one and video two, so total number of VFA scores is 66 scores (n=3\*22). The video stimuli sub-groups were different types of stimuli that were scored either in video one, video two, or both. A child's total VFA score for a given clinical rater and a given video or stimuli group was calculated by summing the rater's scores for the child's response to each visual stimuli in the video or video stimuli group. Statistical significance of the interclass correlation coefficient is reported indicated by \* $p < .05$ and + $p < .10$ .

There were six participants who were not able to complete Teller Acuity Cards II testing and who had not been able to complete any in-clinic acuity testing in the past year. Four of these children did have a present Fix-and-Follow (FF) documented in their medical history and two children had an absent FF. Both children with an absent FF were scored by the VFA (total fixation scores of 0.555 and 0.269).

Table 2 also shows the intraclass correlation coefficients (ICCs) between the three raters for Video One, Video Two, and Video One and Two combined, as well as the sub-grouped attributes of interest (fixation and smooth pursuits, and white and black background). All ICCs for Video One, Video Two, and Video One and Two combined demonstrated good reliability. (i.e., where good reliability^27^ is defined as an ICC of 0.75-0.90^27^) with confidence intervals showing reasonable precision (see Table 2).

The ICC for the smooth pursuits sub-group was 0.679 (95% Confidence Interval (CI): 0.478, 0.830) whereas the fixation subgroup had good agreement with an ICC of 0.848 (95% CI: 0.723, 0.923). There were no apparent differences between the ICCs of the black background and white background subgroups, and these ICCs were also similar to the general video groups. It was not possible to obtain ICCs for Video Three, due to the frequency of zero scores across multiple participants. Using data visualization of Video Three final scores across the raters there appears to be good agreement in deriving an acuity estimate and strong agreement in determining when a child did not perceive a grating (i.e., cases where a “n/t” was given; See Appendix B),.

There was a strong inverse correlation between clinical acuity and average VFA scores for Video One & Two (Figure 2), and Video One (Video One & Two r = -0.706, p = 0.002, Video One r = -0.701, p = 0.003). There was a moderate inverse correlation with Video Two (Video Two r = -0.678, p = 0.004). The linear regression models for Video One, Video Two, and Video One and Two combined all show an inverse relationship between clinical acuity measures and average VFA scores, with clinical acuity explaining 49.8% of the variability in mean VFA scores for Videos One and Two combined, 49.1% of the variability in mean VFA scores for Video One, and 45.9% of the variability in mean VFA scores for Video Two.

**Figure 2.**
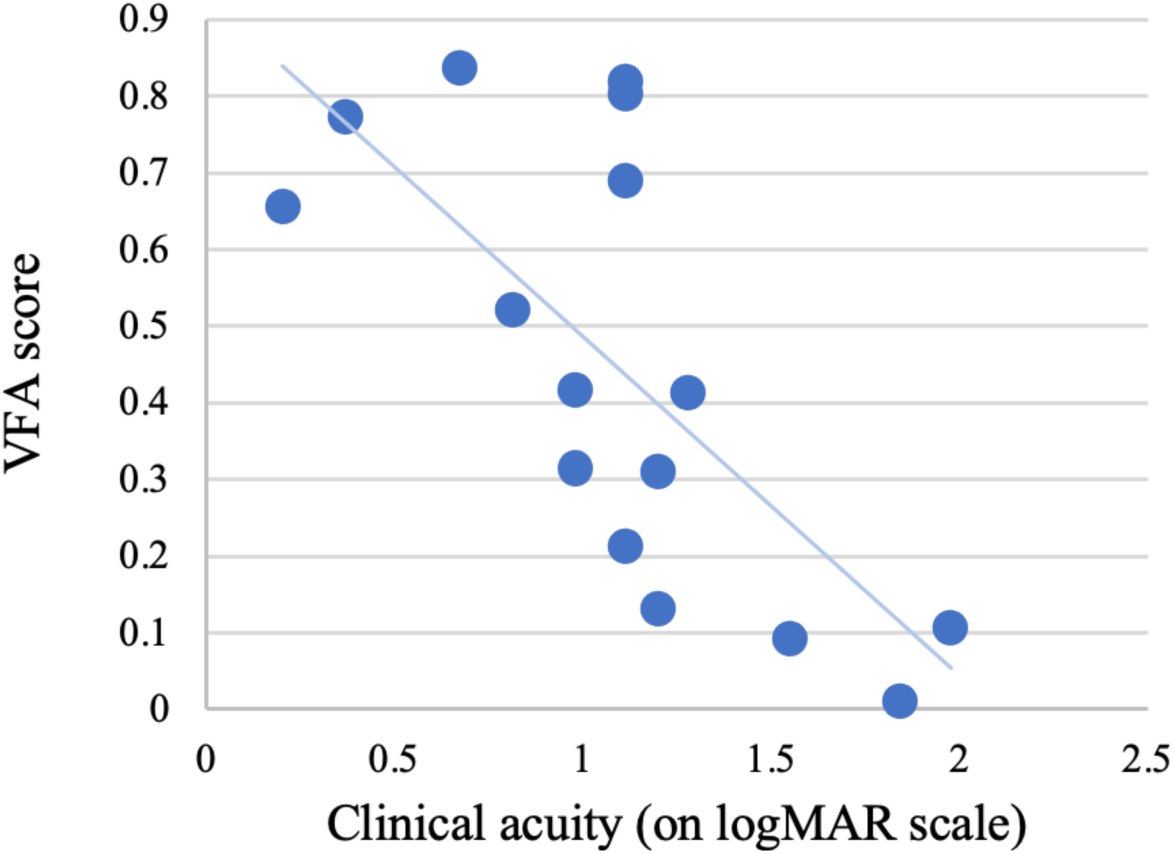
Correlation between children’s VFA scores, clinical acuity, and cognitive age equivalent.

There was a moderate correlation between cognitive age equivalent and average VFA scores (Video One & Two r = 0.518, p = 0.016, Video One r = 0.510, p = 0.018, Video Two r = 0.503, p = 0.020). The most pronounced variability in VFA scores occurred among those with the lowest cognitive age equivalent (less than 10 months). Correlation and regression models were also run for the video sub-groups (fixation, smooth pursuits, black background, white background), and as expected the results were similar to those that we found with the video groups.

There was a strong inverse correlation between cognitive age equivalency and acuity (r = -0.663, p = 0.005) so these were not run together in the regression models. Average acuity estimates from Video Three were moderately correlated with clinical acuity measures (r = -0.606, p = 0.013).

## DISCUSSION

We found good inter-rater agreement for VFA scores, with confidence intervals showing reasonable precision. Our levels of agreement were similar to the Kooiker et al. study which used video-recordings of children with vision impairment to derive quantitative measures of visual function^21^. A similar level of agreement was also found in a study examining inter-rater reliability of visual assessments conducted with children with hemiplegic Cerebral Palsy.^29^

Inter-rater agreement was somewhat weaker for VFA scores of Video One versus Video Two; however, differences in the mean VFA scores were not statistically significant. Differences in attributes of the videos may have influenced the child’s looking behavior in a way that influenced scoring and subsequently agreement. The small differences in agreement may also be due to order effects or to differences in the video attributes.

There was also good agreement among raters for most sub-groupings of video attributes (fixation, black background, and white background) but it was notable that for smooth pursuit stimuli, there was only moderate agreement. Fixation is a visual skill that is typically developed within the first weeks after birth while smooth pursuit typically develops around 2-6 months,^30,31^ and prematurity likely delays the development of smooth pursuit.^32^ Given the clinical profile of the participants, it is reasonable to expect higher scores for fixation than smooth pursuit. The difference in agreement is not as intuitive but may be explained by the fact that impairment in performing smooth pursuit can result in more varied, inaccurate looking behavior that was harder to score. Smooth pursuit required more interpretation by the raters than fixation, and the raters may have interpreted eye movements in the direction of the stimulus differently. Given the high incidence of impairment in smooth pursuit, it is encouraging that the raters still demonstrated moderate inter-rater reliability in rating smooth pursuit stimuli.

When comparing VFA scores to clinically measured acuity, it was expected that there would be a strong inverse correlation between VFA scores and clinically obtained acuity. An inverse correlation was expected because, on the logMAR scale, higher scores indicate poorer acuity. We also expected that the linear regression model would demonstrate a relationship between VFA and acuity, but with a degree of variability, because it is known that the pediatric CVI population typically shows heterogeneity in the level of impairment across different visual functions.^33^ Variability in acuity of babies and young children is also typical.^34^

It was reasonable to find that the relationship between VFA scores and cognitive age equivalents was limited because the assessment was designed to have a low cognitive demand in the areas of attention and competing stimuli. The broad variability in the VFA scores for participants below a 10-month cognitive age equivalent was notable. It would not be unusual for children with CVI, at a similar age equivalence, to have variability in their vision skills.^35^ Future studies examining the construct validity of the VFA will help confirm whether the VFA does in fact offer exciting new specificity in quantifying the visual function of young and medically complex children with CVI.

There was not a strong correlation between Video Three mean acuity estimates and clinical acuity, as we expected given the abbreviated design of Video Three. Our purpose with Video Three was to trial the presentation of gratings by video to this population to see if it may be possible to reliably score acuity in these children from video recording. The high number of zero scores was not overly surprising given low level of visual function across the study population. And, despite the high incidence of zero scores for Video Three, from data visualization there appeared to be reasonable agreement between raters, supporting the addition of a fully calibrated grating acuity assessment into the protocol of future studies using the VFA or a similar video-based assessment.

There were several limitations to this study that we were aware of during the design, and some which later emerged. We understood that using a clinical convenience sample would likely result in a limited spectrum of visual ability among our population. However, we felt this was acceptable for this first proof-of-concept study and would serve our purpose in examining whether the VFA can reliably assess these traditionally difficult to assess children. We also have a limited number of clinical measures for which we can compare the VFA, and this was done to minimize the in-clinic time burden on the families. Based upon these findings, and the enthusiasm caregivers expressed for participating in research for CVI, there is good support for conducting larger follow-up studies with the VFA. Future studies should assess the construct validity and intra-rater reliability under a design that draws from a larger population and which controls for potentially confounding elements such as order effects.

It was anticipated that we would not obtain clinical acuity measures on all children because of low levels of visual function in our sample. But it may also be result of the day to day variability in vision skills that is common for children with CVI. ^36^ Remote assessment could remedy this by capturing windows of optimal visual ability. We can’t be certain which of these factors was of the greatest influence for this study but it did leave us with a group of six participants that were not able to attend or perceive the gratings on the day of assessment. Future designs targeting this population should factor this into the analysis for targeted sample size.

## CONCLUSION

The VFA was shown to be successful in deriving a quantitative score of visual function for young and medically complex children with CVI. This small study found that the VFA has strong inter-rater reliability when used by clinicians specializing in vision. This study also demonstrates the success of a video-based assessment modality for young and medically complex children with CVI, and the utility of examining their visual function from video recording. This method of assessment from video recording could also offer the possibility of capturing video recordings in the home, on a day when the caregiver perceives the child’s visual ability to be at their best.

This would be a helpful option for children with CVI whose vision can vary day-to-day and could help to gather a more accurate assessment of the child’s best visual ability.

## Data Availability

All data produced in the present study are available upon reasonable request to the authors

---

**Appendix A.** VFA Scoring Criteria.

| Score | Definition | Description |
| --- | --- | --- |
| 0 | No visual interest.<br>Fixation or tracking not observed | Child is looking away from the screen, holding hands over face or otherwise not attending to the screen |
| 1 | Glanced in general direction of stimulus | Child is looking at the screen but does not show any indication of fixating or tracking the stimulus. They may move their glance in the general direction of the stimulus such that they may be responding to the presentation of the light or color, but there is no indication of purposefully targeting the stimulus. |
| 2 | Possible fixation/ tracking, but very brief, not convinced | Child shows moments where they appear to accurately fixate or track the stimulus, but it is very brief. Given the brief nature it is difficult to be convinced of their success in locating the stimulus, but given the directionality of their viewing it is possible they successfully fixated or tracked the stimulus very briefly. |
| 3 | Intentional fixation/ tracking, but not sustained | Child shows discrete and accurate fixation or tracking of the stimulus. This may occur one time, or they may accurately return to the stimulus more than one time. |
| 4 | Accurate, short fixation/tracking | Child shows discrete and accurate fixation of the stimulus and holds fixation or tracking for a sustained period of time. Their gaze may break, but when they return to the screen, they are able to achieve accurate fixation on the stimulus. For a child with latency, they may often perform at this level because they require extra time to obtain fixation. |
| 5 | Steady, mostly sustained fixation/tracking | Child shows discrete and accurate fixation of the stimulus and hold fixation or tracking the majority of the time that the stimulus is presented. They may break attention to the stimulus briefly, but they return to screen and immediately achieve accurate fixation on the stimulus. |

**Appendix B.** Total visual acuity estimates for children assessed from three presentations of three different grating stimuli presented in Video Three.

| Participant Number | Total VFA Score: Rater One | Total VFA Score: Rater Two | Total VFA Score: Rater Three | Mean of Total VFA Scores |
| --- | --- | --- | --- | --- |
| 1 | n/t | n/t | n/t | n/t |
| 2 | n/t | n/t | n/t | n/t |
| 3 | 0.67 | 0.67 | 0.67 | 0.67 |
| 4 | n/t | 1.28 | 1.85 | 1.04 |
| 5 | 0.67 | 1.85 | 0.67 | 1.06 |
| 6 | n/t | n/t | n/t | n/t |
| 7 | 1.85 | 1.85 | 0.67 | 1.45 |
| 8 | 0.67 | 0.67 | n/t | 0.45 |
| 9 | n/t | n/t | n/t | n/t |
| 10 | 1.28 | 1.28 | 0.67 | 1.08 |
| 11 | n/t | n/t | 1.85 | 0.62 |
| 12 | n/t | n/t | n/t | n/t |
| 13 | n/t | n/t | n/t | n/t |
| 14 | 1.85 | 1.85 | n/t | 1.23 |
| 15 | 0.67 | 0.67 | 0.67 | 0.67 |
| 16 | n/t | n/t | n/t | n/t |
| 17 | n/t | n/t | n/t | n/t |
| 18 | 1.85 | 1.85 | 1.28 | 1.66 |
| 19 | 1.85 | n/t | n/t | 0.62 |
| 20 | n/t | n/t | n/t | n/t |
| 21 | n/t | n/t | n/t | n/t |
| 22 | 1.85 | 1.85 | 1.28 | 1.66 |

Note: All ratings are reported as the logarithm of the minimum angle of resolution (logMAR) with exception of those that could not be rated which were given a rating of “n/t”.

## REFERENCES

1. Salt A, Sargent J. Common visual problems in children with disability. Archives of disease in childhood. 2014;99(12):1163–1168.

2. Ameenat Lola S, Lucinda T, Jugnoo R. Epidemiology of blindness in children. Archives of Disease in Childhood. 2017;102(9):853. doi:10.1136/archdischild-2016-310532

3. Sakki HE, Dale NJ, Sargent J, Perez-Roche T, Bowman R. Is there consensus in defining childhood cerebral visual impairment? A systematic review of terminology and definitions. British Journal of Ophthalmology. 2018;102(4):424–432.

4. Philip SS, Dutton GN. Identifying and characterising cerebral visual impairment in children: a review. Clin Exp Optom. May 2014;97(3):196–208. doi:10.1111/cxo.12155

5. Lam FC, Lovett F, Dutton GN. Cerebral visual impairment in children: a longitudinal case study of functional outcomes beyond the visual acuities. Journal of Visual Impairment & Blindness. 2010;104(10):625–635.

6. Ospina LH. Cortical visual impairment. Pediatr Rev. Nov 2009;30(11):e81–90. doi:10.1542/pir.30-11-e81

7. Jimenez-Gomez A, Fisher KS, Zhang KX, Liu C, Sun Q, Shah VS. Longitudinal neurological analysis of moderate and severe pediatric cerebral visual impairment. Frontiers in Human Neuroscience. 2022;16:772353.

8. Handa S, Saffari SE, Borchert M. Factors associated with lack of vision improvement in children with cortical visual impairment. Journal of Neuro-ophthalmology. 2018;38(4):429–433.

9. Chorna OD, Guzzetta A, Maitre NL. Vision Assessments and Interventions for Infants 0-2 Years at High Risk for Cerebral Palsy: A Systematic Review. Pediatr Neurol. Nov 2017;76:3–13. doi:10.1016/j.pediatrneurol.2017.07.011

10. Morelli F, Aprile G, Martolini C, et al. Visual Function and Neuropsychological Profile in Children with Cerebral Visual Impairment. Children (Basel). Jun 19 2022;9(6) doi:10.3390/children9060921

11. Manley CE, Walter K, Micheletti S, et al. Object identification in cerebral visual impairment characterized by gaze behavior and image saliency analysis. Brain and Development. 2023/05/13/ 2023;10.1016/j.braindev.2023.05.001

12. Chang MY, Borchert MS. Methods of visual assessment in children with cortical visual impairment. Current Opinion in Neurology. 2021;34(1):89–96.

13. Dutton G, Ballantyne J, Boyd G, et al. Cortical visual dysfunction in children: a clinical study. Eye. 1996;10(3):302–309.

14. Pamir Z, Bauer CM, Bailin ES, Bex PJ, Somers DC, Merabet LB. Neural correlates associated with impaired global motion perception in cerebral visual impairment (CVI). NeuroImage: Clinical. 2021/01/01/ 2021;32:102821. 10.1016/j.nicl.2021.102821

15. Altınbay D, Taşkın İ. Evaluation of Behavioral Characteristics in Response to Visual Stimuli in Infants with Cerebral Visual Impairment. Turkish journal of ophthalmology. 2023;53(1):1–7.

16. Mooney SWJ, Alam NM, Prusky GT. Tracking-Based Interactive Assessment of Saccades, Pursuits, Visual Field, and Contrast Sensitivity in Children With Brain Injury. Front Hum Neurosci. 2021;15:737409. doi:10.3389/fnhum.2021.737409

17. Ortibus E, Lagae L, Casteels I, Demaerel P, Stiers P. Assessment of cerebral visual impairment with the L94 visual perceptual battery: clinical value and correlation with MRI findings. Developmental Medicine & Child Neurology. 2009;51(3):209–217.

18. Itzhak NB, Kooiker M, Pel J, Ortibus E. Including visual orienting functions into cerebral visual impairment screening: Reliability, variability, and ecological validity. Research in Developmental Disabilities. 2023;132:104391.

19. Chang MY, Borchert MS. Validity and reliability of eye tracking for visual acuity assessment in children with cortical visual impairment. Journal of American Association for Pediatric Ophthalmology and Strabismus. 2021;25(6):334. e1–334. e5.

20. Almagati R, Kran BS. Implications of a Remote Study of Children With Cerebral Visual Impairment for Conducting Virtual Pediatric Eye Care Research: Virtual Assessment Is Possible for Children With CVI. Brief Research Report. Frontiers in Human Neuroscience. 2021-September-14 2021;15doi:10.3389/fnhum.2021.733179

21. Kooiker MJ, Pel JJ, Verbunt HJ, de Wit GC, van Genderen MM, van der Steen J. Quantification of visual function assessment using remote eye tracking in children: validity and applicability. Acta ophthalmologica. 2016;94(6):599–608.

22. Tanke N, Barsingerhorn AD, Goossens J, Boonstra FN. The Developmental Eye Movement Test as a Diagnostic Aid in Cerebral Visual Impairment. Front Hum Neurosci. 2021;15:732927. doi:10.3389/fnhum.2021.732927

23. Pel JJ, Dudink J, Vonk M, Plaisier A, Reiss IK, van der Steen J. Early identification of cerebral visual impairments in infants born extremely preterm. Developmental Medicine & Child Neurology. 2016;58(10):1030–1035.

24. 24. Bayley N. Bayley scales of infant and toddler development. 2006;

25. 25. Teller DD, Velma; Mayer, Luisa. Teller Acuity Cards TAC II Reference and Instruction Manual. Precision Vision; 2014. https://storage.googleapis.com/stateless-precision-vision/2020/08/f9e53b60-tacii-manual_facing-pages_rev08312020.pdf

26. Nichols DS, Case-Smith J. Reliability and validity of the Pediatric Evaluation of Disability Inventory. Pediatric Physical Therapy. 1996;8(1):15–24.

27. Koo TK, Li MY. A guideline of selecting and reporting intraclass correlation coefficients for reliability research. Journal of chiropractic medicine. 2016;15(2):155–163.

28. Harris PA, Taylor R, Thielke R, Payne J, Gonzalez N, Conde JG. Research electronic data capture (REDCap)—A metadata-driven methodology and workflow process for providing translational research informatics support. Journal of Biomedical Informatics. 2009/04/01/ 2009;42(2):377-381. 10.1016/j.jbi.2008.08.010

29. Auld M, Boyd R, Moseley GL, Johnston L. Seeing the gaps: a systematic review of visual perception tools for children with hemiplegia. Disability and Rehabilitation. 2011/01/01 2011;33(19-20):1854-1865. doi:10.3109/09638288.2010.549896

30. Krieber-Tomantschger M, Pokorny FB, Krieber-Tomantschger I, et al. The development of visual attention in early infancy: Insights from a free-viewing paradigm. Infancy. 2022;27(2):433–458.

31. Christina P, Frank P, Irene G. Smooth pursuit in infants: maturation and the influence of stimulation. British Journal of Ophthalmology. 2012;96(1):73. doi:10.1136/bjo.2010.191726

32. Strand Brodd K, Grönqvist H, Holmström G, Grönqvist E, Rosander K, Ewald U. Development of smooth pursuit eye movements in very preterm born infants: 3. Association with perinatal risk factors. 10.1111/j.1651-2227.2011.02449.x. Acta Paediatrica. 2012/02/01 2012;101(2):164-171. 10.1111/j.1651-2227.2011.02449.x

33. Chandna A, Ghahghaei S, Foster S, Kumar R. Higher visual function deficits in children with cerebral visual impairment and good visual acuity. Frontiers In Human Neuroscience. 2021;15:711873.

34. Leat SJ, Yadav NK, Irving EL. Development of visual acuity and contrast sensitivity in children. Journal of optometry. 2009;2(1):19–26.

35. Ortibus E, Fazzi E, Dale N. Cerebral Visual Impairment and Clinical Assessment: The European Perspective. Seminars in Pediatric Neurology. 2019/10/01/ 2019;31:15-24. 10.1016/j.spen.2019.05.004

36. Dutton GN. The spectrum of cerebral visual impairment as a sequel to premature birth: an overview. Documenta Ophthalmologica. 2013/08/01 2013;127(1):69-78. doi:10.1007/s10633-013-9382-1

